# Excess deaths in China during SARS-CoV-2 viral waves in 2022-2023

**DOI:** 10.1101/2023.10.18.23297204

**Authors:** Prabhat Jha, Patrick E Brown, Teresa Lam, Ed Morawski, Angus Reid

## Abstract

**Background:** The extent to which the Omicron variant of SARS-CoV-2 raised death rates in China during its viral wave of December 2022-January 2023 remains undocumented.

**Methods:** We worked with an established national survey organization to survey 8,004 adults in all 31 administrative areas of China to ask about deaths in families since January 2020. We examined agespecific death rates, focusing on deaths above age 60 years, and at 15-59 years. We compared these to the United Nations (UN) estimates of age-specific mortality in 2019.

**Findings:** The survey participants were broadly similar to the 2020 census and other national surveys in age, sex, region, and smoking status, but had lower SARS-CoV-2 vaccination rates and higher education levels. There were no differences between reporting of deaths during the Omicron period versus earlier. The survey captured 456 deaths, of which 329 occurred at ages 60+ years and 212 were women. At ages 60+ years, death rates per 1000 rose 242% (95%CI 128-398%) during December 2022-January 2023. Deaths at ages 15-59 years did not rise appreciably. The UN estimates approximately 675,000 deaths per month at ages 60+ years in 2019. If rates doubled nationally as in our survey, China had approximately 1·35 million excess deaths over the two months.

**Interpretation:** China experienced a sharp but short increase in excess deaths among its elderly during the Omicron wave. If death registry data corroborate our estimates of substantial excess deaths in China, the worldwide estimates of excess deaths to 2023 may need upward adjustment.

**Funding:** Canadian Institutes of Health Research

## Introduction

The People’s Republic of China adopted stringent lockdowns, quarantine, mass testing, and vaccination to combat the SARS-CoV-2 pandemic.^1,2^ Despite some early success,^2^ local outbreaks occurred and a national viral wave of over 100,000 test-confirmed cases ensued in March-April 2022. After abandoning most restrictions and the arrival of the Omicron variants, China reported a sharp increase in cases in December 2022-January 2023.^1^ Over 100,000 PCR test-confirmed COVID deaths occurred in hospitals during this period.^1^ However, many deaths, even in hospital, went untested or occurred at home.

Excess mortality from all causes is a robust method to capture the direct and indirect mortality from SARS-CoV-2 viral waves.^3,4^ Direct estimates of excess mortality have not been reported after the original viral wave in Spring 2020.^2^ We calculated excess mortality in a nationally representative survey of China.

## Methods

### Recruitment

We worked with an established survey research organization with an online sampling platform. The survey research organization has over two decades of experience in Asia, including in China. The online platform recruits members from mainland China, who receive points used for various merchandise from participation in each survey. Overall internet penetration exceeds 75% in China.^5^ As of April 2023, the panel covered over three million adults. Additional details of the recruitment channels and quality control are provided in the Appendix. The study received ethical approval by the Unity Health Toronto Research Ethics Board (REB # 15-231).

### Sampling

For this study, the sampling strategy involved creating a profile that was representative of China’s four regions (Eastern, Central, Western, and Northeastern) and broadly by age group (18-59 and 60+) and sex. Additionally, the sample frame was balanced on five tiers that represented over 700 cities, and several income ranges with lower income participants.

### Survey

The survey focused on changes in consumption, travel, and household structure during the COVID-19 pandemic, to understand the economic recovery. We developed an online survey with 38 items in February 2023, and pretested it among 200 participants in March 2023, with the final survey implemented in early April 2023. We compared our respondents’ characteristics with Census 2020 or nationally representative surveys on income and health behavior (smoking, SARS-CoV-2 vaccination).^6-11^ Additionally, our participants answered questions about the time (month and year) and age for each death that occurred in their families since Lunar New Year (January 25) 2020. Participants took about 8-12 minutes to complete the survey.

### Analyses

Any deaths in participants’ families were captured by a dichotomous variable (1=any death, 0=no death), and a discrete variable tallied the number of deaths. For participants with at least one death in the family since Lunar New Year (January 25) 2020, another dichotomous variable (1=since November 2021, 0=earlier) captured deaths that occurred after the arrival of the Omicron variant.

We smoothed the study’s monthly death rates using 3-month rolling averages (although 2-month and 4-month averages showed similar patterns; data not shown). The numerator of the monthly death rates was the number of deaths by age group by month. The denominator was the sum of all family members by age group and month (Appendix 2).

We used China’s population and deaths reported on the World Population Prospects 2022 by the United Nations Population Division to calculate the national age-specific death rates from 2012-2019.^5^ We divided the annual rates by 12 to obtain the average monthly rates. We used logistic regression to determine the likelihood of having a death in the family vs. not, and of having any death in the family since November 2021 vs. any earlier death since Lunar New Year (January 25) 2020. We used Poisson regression, with appropriate confidence intervals, to estimate changes in the age-specific survey death rates during Dec 2022-Jan 2023 compared to earlier periods. We used Stata/MP 17 to conduct all analyses.^12^ Our data and codebook are available on GitHub: https://github.com/ChinaStudy2023.

## Results

About 210,000 panelists were invited, of whom 8,004 completed the full survey. The response rate of about 4% is typical of comparable online surveys in China. Our sample was broadly representative of the regional population distribution of China’s 31 administrative regions (Table 1). Compared to the 2020 census and other national surveys, study participants were broadly representative by age, sex, region, and smoking status, but had lower SARS-CoV-2 vaccination rates and higher education levels. Households reporting a death differed little from those that did not, except for more deaths occurring in households with a smoker and fewer deaths in more educated households. There were no differences between reporting of deaths during the Omicron period versus earlier.

**Table 1.** Characteristics of survey participants compared to national census/other data and comparison of households with and without a death.

| Variable | Proportions<br>in<br>Census/other<br>surveys | Study number (N) and proportions |  |  | Survey-weighted odds ratio (OR) in logistic regression |  |  |  |
| --- | --- | --- | --- | --- | --- | --- | --- | --- |
|  |  | Unweighted<br>N | Weighted % | Weighted % with a<br>death in the family | Any death in family versus<br>(n=354) versus no deaths<br>(n=7650) |  | Deaths in Nov 2021 or<br>after (n=144) versus deaths<br>earlier (n=210) |  |
|  |  |  |  |  | OR | (95% CI) | OR | (95% CI) |
| **Region <sup>a</sup>** |  |  |  |  |  |  |  |  |
| Eastern | 40% | 3,204 | 40% | 35% | Reference |  | Reference |  |
| Central | 26% | 2,048 | 26% | 26% | 1.2 | (0.7 - 2.1) | 1.3 | (1.1 - 1.7) |
| Western | 27% | 2,179 | 27% | 33% | 1.4 | (1.0 - 2.1) | 1.2 | (0.6 - 2.4) |
| Northeastern | 7% | 573 | 7% | 6% | 0.9 | (0.4 - 2.0) | 2.3 | (0.3 - 17.4) |
| **Age group <sup>b</sup>** |  |  |  |  |  |  |  |  |
| Age 15-59 | 78% | 7099 | 86% | 82% | Reference |  | Reference |  |
| Age 60 or older | 22% | 905 | 14% | 18% | 1.3 | (0.7 - 2.3) | 0.8 | (0.3 - 2.1) |
| **Sex <sup>c</sup>** |  |  |  |  |  |  |  |  |
| Male | 51% | 4,056 | 50% | 48% | Reference |  | Reference |  |
| Female | 49% | 3,945 | 50% | 52% | 1.4 | (1.1 - 1.9) | 0.6 | (0.3 - 1.1) |
| **Education level <sup>d</sup>** |  |  |  |  |  |  |  |  |
| Primary school | 25% | 299 | 4% | 10% | Reference |  | Reference |  |
| Secondary school | 15% | 1,374 | 18% | 19% | 0.4 | (0.3 - 0.6) | 1.1 | (0.3 - 4.7) |
| Some university/Vocational school |  | 2,459 | 31% | 29% | 0.4 | (0.2 - 0.7) | 1.1 | (0.2 - 5.3) |
| University | 19% | 3,863 | 47% | 41% | 0.4 | (0.2 - 0.8) | 0.9 | (0.2 - 3.6) |
| Prefer not to answer |  | 9 |  |  |  |  |  |  |
| **Monthly pre-tax household income** |  |  |  |  |  |  |  |  |
| 5000 RMB or less |  | 3904 | 49% | 48% | Reference |  | Reference |  |
| 5001 RMB or more |  | 4077 | 51% | 52% | 1.2 | (1.0 - 1.4) | 0.9 | (0.6 - 1.2) |
| Prefer not to answer |  | 23 |  |  |  |  |  |  |
| **Smoking history <sup>e</sup>** |  |  |  |  |  |  |  |  |
| Never |  | 5,585 | 70% | 57% | Reference |  | Reference |  |
| Ever | 31% | 2,419 | 30% | 43% | 2.1 | (1.5 - 2.9) | 0.7 | (0.3 - 1.4) |
| **SARS-CoV-2 vaccinated versus not <sup>f</sup>** | 89% | 6,284 | 78% | 80% | 1.1 | (0.6 - 2.0) | 0.7 | (0.4 - 1.4) |
| **Number of deaths reported in family** |  |  |  |  |  |  |  |  |
| None |  | 7,650 | 96% |  |  |  |  |  |
| One |  | 285 | 4% | 81% |  |  |  |  |
| Two or more |  | 51 | 1% | 19% |  |  |  |  |

NOTES: RMB=Renminbi; One respondent who preferred not to describe their sex and two who preferred not to report their education are excluded from the second logistic regression; See Appendix for References: a. Communiqué of the Seventh National Population Census (No. 3)-Population by Region. National Bureau of Statistics of China, 2021; b. International Database: World Population Estimates and Projections. United States Census Bureau; c. China's latest census reports more balanced gender ratio. Xinhuanet, 2021; d. Communiqué of the Seventh National Population Census (No. 6)-Education Attainment. National Bureau of Statistics of China, 2021; e. Zhang M, Yang L, Wang L, et al. Trends in smoking prevalence in urban and rural China, 2007 to 2018: Findings from 5 consecutive nationally representative cross-sectional surveys. PLoS Med 2022; f. Coronavirus (COVID-19) Vaccinations. OurWorldInData.org, 2022.

The survey captured 456 deaths, of which 329 occurred at 60+ years and 212 were women. At 60+ years, the average death rates per 1000 fluctuated but rose 242% (95%CI 128-398%) during December 2022-January 2023 (Figure 1). Survey death rates at these ages were lower than the UN death rates, which are more complete as they draw upon death registration and census data. Death rates at ages 15-59 showed less fluctuation and were closer to the UN death rates.

**Figure 1.**
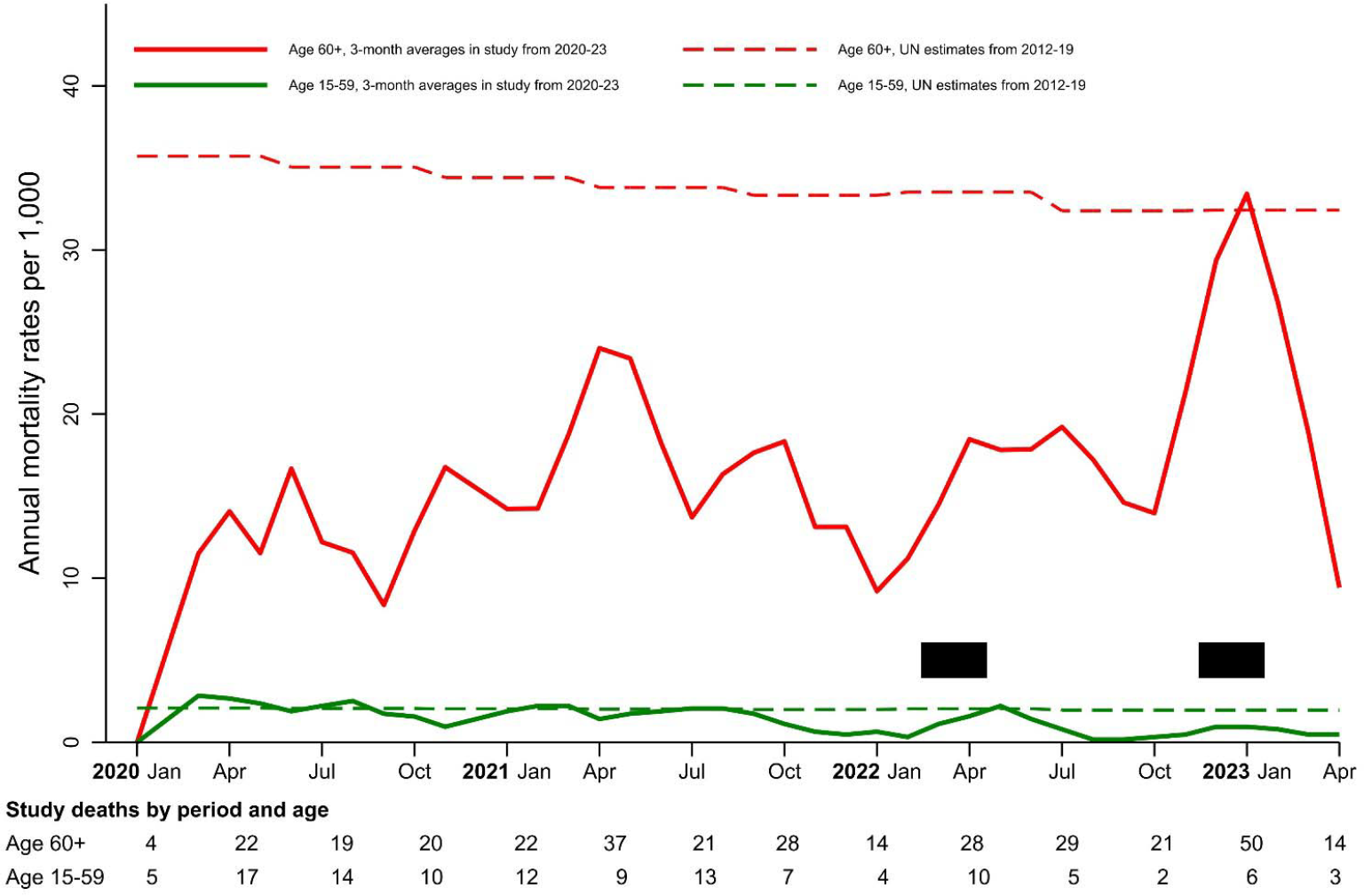
Mortality rates per 1000 at ages 60 years or older and 15-59 years in a nationally representative survey in China from 2020-23 compared to United Nations estimates for earlier years. Peak viral periods in March-April 2022 and Dec 2022-Jan 2023^1^ are shown in the black rectangles. The numbers of study deaths are shown in the text below the figure. Study deaths use rolling three-month averages. The annual mortality rates are per 1,000 person-years. The smoothed UN estimated deaths are for 2012-19 (https://population.un.org/wpp/).

Using the UN estimates, China had about 8·1 million deaths at ages 60+ in 2019,^5^ which (ignoring seasonality) equates to approximately 675,000 deaths monthly. If death rates doubled nationally during December 2022-January 2023, as they clearly did so in our survey, mainland China had 1·35 million excess deaths at 60+ years.

## Discussion

Our estimate of 1·35 million excess deaths in China’s last SARS-CoV-2 wave is crude by necessity, but consistent results drawn on modelling based on increases in cremations^13^ and death registration data in China’s special administrative regions. Hong Kong saw about 70% excess deaths in March-April 2022 (when the increase in cases was smaller than in December 2022-January 2023) and Macau saw about 170% excess deaths during the later peak.^14^ Both regions have better health systems and reporting than the whole of China. Our estimate is lower than that based on internet searches for funerals.^15^ Excess deaths were concentrated at ages 60+ also during the original wave of spring 2020.^2^ Additionally, China has high overall vaccination coverage of 90%, but its elderly are disproportionally less fully (3-doses) vaccinated.^16^

We did not observe over-reporting of deaths during the Omicron wave versus earlier. Indeed, we sequenced questions to avoid such spurious reporting. Nevertheless, sample surveys generally underestimate deaths^17^ and are no substitute for timely release of China’s high-quality death registration data. China’s 2025 Census and its Disease Surveillance Point system (covering 20% of the population) could each quantify excess deaths directly by reporting deaths from January 2020 onward by location, age, sex, and date.

Despite an older population, China’s excess deaths during the Omicron wave of December 2022-January 2023 were about half of what India experienced during its very large Delta wave of April-May 2021.^2-4^ If indeed China had substantial excess deaths in late 2022/early 2023, they would suggest a significant upward revision of worldwide excess deaths from SARS-CoV-2 infection through mid-2023 from the estimated 13-17 million deaths as of December 2021.^3^

## Data Availability

Our data and codebook are available on GitHub: https://github.com/ChinaStudy2023.

## Acknowledgements

We thank Zhengming Chen with advice on the survey design, Fatima LaHay for advice on the sampling panel, and Leslie Newcombe for editorial assistance. This research is supported by the Canadian Institutes of Health Research.

## Appendix 1 Recruitment and quality control

Recruitment channels include working with affiliates, using banners at commercial centres to recruit specific demographics (e.g., younger adults buying vehicles), relying on word of mouth from existing members, and using social media platforms with careful checks to counter robotic requests to register. The organization removed any subject suspected of being fraudulent (or robotic). The survey research organization conducts various quality control steps to manage the panel, including verification of IP addresses, devices’ information, and subjects’ area of residence, as well as validity of emails and phone numbers. New panellists receive at least one quality-control check every month and receive at least two requests to update their demographic information each year. Panellists who are unresponsive for more than one year are removed. Additional internal data quality checks, including detection for artificial intelligence, are also performed on each panel.

## Appendix 2 Supplementary information for calculating family size

Each participant’s family size was defined as the sum of participants themselves, a living partner, a living mother, a living father, the number of living brothers, sisters, sons, and daughters, as well as the number of new members that joined their families since Lunar New Year 2020. Then, because we only asked for ages for the first three brothers, first three sisters, first three sons, and first three daughters, we made assumptions regarding the ages of subsequent siblings and offspring. For brothers and sisters, we used the participants’ age group for the ones with unknown ages. For sons and daughters, because we asked for ages of “first son”, “second son”, and so forth, we assumed that people responded from the oldest to the youngest. Then, any son or daughter beyond the first three used the same age group as the third one. Lastly, for all months after January 2020, family size was calculated as the difference between family size and deaths in the previous month. This yielded larger family sizes than observed in the 2020 census,^1,2^ which would result in lower death rates, given a larger denominator of living adults. However, these lower death rates were observed consistently at both ages 60 or older and below these ages. As a further check on the denominators and given the study sample size (n=8,004) is a small fraction of China’s population in 2020 Census, we applied the same fraction to obtain age-specific population sizes for the age-specific mortality rates. This method yielded higher death rates, but no change in the peak and non-peak distribution of deaths (data not shown).

## Appendix 3 STROBE Statement

|  | Item No. | Recommendation | Relevant text from manuscript (page no) |
| --- | --- | --- | --- |
| Title and abstract | 1 | (a) Indicate the study’s design with a commonly used term in the title or the abstract | Excess deaths in China during SARS-CoV-2 viral waves in 2022-2023 (1) |
|  |  | (b) Provide in the abstract an informative and balanced summary of what was done and found | NA |
| Introduction |  |  |  |
| Background/rationale | 2 | Explain the scientific background and rationale for the investigation being reported | After abandoning most restrictions and the arrival of the Omicron variants, China reported a sharp increase in cases in December 2022-January 2023. Over 100,000 PCR test-confirmed COVID deaths occurred in hospitals during this period. However, many deaths, even in hospital, went untested or occurred at home. (3) |
| Objectives | 3 | State specific objectives, including any pre-specified hypotheses | We calculated excess mortality in a nationally representative survey of China. (3) |
| Methods |  |  |  |
| Study design | 4 | Present key elements of study design early in the paper | The survey focused on changes in consumption, travel, and household structure during the COVID-19 pandemic, to understand the economic recovery. We developed an online survey with 38 items in February 2023, and pretested it among 200 participants in March 2023, with the final survey implemented in early April 2023.<br>Additionally, our participants answered questions about the time (month and year) and age for each death that occurred in their families since Lunar New Year (January 25) 2020. Participants took about 8-12 minutes to complete the survey. (4) |
| Setting | 5 | Describe the setting, locations, and relevant dates, including periods of recruitment, exposure, follow-up, and data collection | About 210,000 panelists were invited, of whom 8,004 completed the full survey. (5) |
| Participants | 6 | (a) Give the eligibility criteria, and the sources and methods of selection of participants. Describe methods of follow-up | Recruitment channels include working with affiliates, using banners at commercial centres to recruit specific demographics (e.g., younger adults buying vehicles), relying on word of mouth from existing members, and using social media platforms with careful checks to counter robotic requests to register. (Appendix 1)<br>[...] the sampling strategy involved creating a profile that was representative of China’s<br>four regions (Eastern, Central, Western, and Northeastern) and broadly by age group (18-59 and 60+) and sex. Additionally, the sample frame was balanced on five tiers that represented over 700 cities, and several income ranges with lower income participants. (3) |
|  |  | (b) For matched studies, give matching criteria |  |
| Variables | 7 | Clearly define all outcomes, exposures, predictors, potential confounders, and effect modifiers. Give diagnostic criteria, if applicable | Any deaths in participants' families were captured by a dichotomous variable (1=any death, 0=no death), and a discrete variable tallied the number of deaths. For participants with at least one death in the family since Lunar New Year 2020, another dichotomous variable (1=since November 2021, 0=earlier) captured deaths that occurred after the arrival of the Omicron variant. (4) |
| Data sources/<br>measurement | 8* | For each variable of interest, give sources of data and details of methods of assessment (measurement). Describe comparability of assessment methods if there is more than one group | The numerator of the monthly death rates was the number of deaths by age group by month. The denominator was the sum of all family members by age group and month. (4) |
| Bias | 9 | Describe any efforts to address potential sources of bias | We did not observe over-reporting of deaths during the Omicron wave versus earlier. Indeed, we sequenced questions to avoid such spurious reporting. (6) |
| Study size | 10 | Explain how the study size was arrived at | See No. 5 and No. 6 |
| Quantitative variables | 11 | Explain how quantitative variables were handled in the analyses. | NA |
| Statistical methods | 12 | (a) Describe all statistical methods, including those used to control for confounding | We smoothed the study's monthly death rates using 3-month rolling averages (although 2-month and 4-month averages showed similar patterns). [...] We used logistic regression to determine the likelihood of having a death in the family vs. not, and of having any death in the family since November 2021 vs. any earlier death since Lunar New Year 2020. [...] We used Poisson regression, with appropriate confidence intervals, to estimate changes in the age-specific survey death rates during Dec 2022-Jan 2023 compared to earlier periods. (4) |
|  |  | (b) Describe any methods used to examine subgroups and interactions | NA |
|  |  | (c) Explain missing data | NA |
|  |  | (d) If applicable, explain how loss to follow-up was addressed | NA |
|  |  | (e) Describe any sensitivity analyses | NA |
| **Results** |  |  |  |
| Participants | 13 * | (a) Report numbers of individuals at each stage of study—eg numbers potentially eligible, examined for eligibility, confirmed eligible, included in the study, completing follow-up, and analysed | Table 1 |
|  |  | (b) Give reasons for non-participation at each stage | NA |
|  |  | (c) Consider use of a flow diagram | NA |
| Descriptive data | 14 * | (a) Give characteristics of study participants (eg demographic, clinical, social) and information on exposures and potential confounders | Table 1 |
|  |  | (b) Indicate number of participants with missing data for each variable of interest | NA |
|  |  | (c) Summarise follow-up time (eg, average and total amount) | NA |
| Outcome data | 15 * | Report numbers of outcome events or summary measures over time | Figure 1 |
| Main results | 16 | (a) Give unadjusted estimates and, if applicable, confounder-adjusted estimates and their precision (e.g., 95% confidence interval). | NA |
|  |  | (b) Report category boundaries when continuous variables were categorized | Table 1 |
|  |  | (c) If relevant, consider translating estimates of relative risk into absolute risk | NA |
| Other analyses | 17 | Report other analyses done—e.g. analyses of subgroups and interactions, and sensitivity analyses | NA |
| **Discussion** |  |  |  |
| Key results | 18 | Summarize key results with reference to study objectives | At 60+ years, the average death rates per 1000 fluctuated but rose 242% (95%CI 128-398%) during December 2022-January 2023. [...] Death rates at ages 15-59 showed less fluctuation and were closer to the UN death rates. (5) |
| Limitations | 19 | Discuss limitations of the study, taking into account sources of potential bias or imprecision. Discuss both direction and magnitude of any potential bias | Nevertheless, sample surveys generally underestimate deaths and are no substitute for timely release of China's high-quality death registration data. (6) |
| Interpretation | 20 | Give a cautious overall interpretation of results considering objectives, limitations, multiplicity of analyses, results from similar studies, and other relevant evidence | Despite an older population, China's excess deaths during the Omicron wave of December 2022-January 2023 were about half of what India experienced during its very large Delta wave of April-May 2021. If indeed China had substantial excess deaths in late 2022/early 2023, they would suggest a significant upward revision of worldwide excess deaths from SARS-CoV-2 infection through mid-2023 from the estimated 13-17 million deaths as of December 2021. (6) |
| Generalizability | 21 | Discuss the generalizability (external validity) of the study results | Compared to the 2020 census and other national surveys, study participants were broadly representative by age, sex, region, and smoking status, but had lower SARS-CoV-2 vaccination rates and higher education levels. (5) |
| **Other information** |  |  |  |
| Funding | 22 | Give the source of funding and the role of the funders for the present study and, if applicable, for the original study on which the present article is based | Canadian Institutes of Health Research (2)<br>The funder had no role in data collection, analysis, interpretation of the findings, writing of the paper, or the decision to submit. (5) |

## References

1. China’s COVID-19 Response. 2023. https://en.chinacdc.cn/news/latest/202301/W020230126558725888448.pdf (accessed August 4 2023).

2. Liu J, Zhang L, Yan Y, et al. Excess mortality in Wuhan city and other parts of China during the three months of the covid-19 outbreak: findings from nationwide mortality registries. BMJ 2021; 372: 415.

3. Msemburi W, Karlinsky A, Knutson V, Aleshin-Guendel S, Chatterji S, Wakefield J. The WHO estimates of excess mortality associated with the COVID-19 pandemic. Nature 2023; 613(7942): 130–7.

4. Jha P, Deshmukh Y, Tumbe C, et al. COVID mortality in India: National survey data and health facility deaths. Science 2022; 375(6581): 667–71.

5. The State Council of the People’s Republic of China. China’s internet users total 1.079 bln with vibrant digital infrastructure, internet app growth. August 28, 2023. https://english.www.gov.cn/archive/statistics/202308/28/content_WS64ec5126c6d0868f4e8dee21.html (accessed October 11 2023).

6. Zhang M, Yang L, Wang L, et al. Trends in smoking prevalence in urban and rural China, 2007 to 2018: Findings from 5 consecutive nationally representative cross-sectional surveys. PLoS Med 2022; 19(8): e1004064.

7. United States Census Bureau. International Database: World Population Estimates and Projections. December 21, 2021. https://www.census.gov/programs-surveys/international-programs/about/idb.html (accessed August 1 2023).

8. China’s latest census reports more balanced gender ratio. 2021. http://www.xinhuanet.com/english/2021-05/11/c_139938390.htm (accessed August 1 2023).

9. Communiqué of the Seventh National Population Census (No. 6)-Education Attainment. May 11, 2021 2021. http://www.stats.gov.cn/english/PressRelease/202105/t20210510_1817191.html (accessed August 1 2023).

10. Mathieu E, Ritchie H, Rodes-Guirao L, et al. Coronavirus (COVID-19) Vaccinations. 2020. https://ourworldindata.org/covid-vaccinations (accessed August 1 2023).

11. Communiqué of the Seventh National Population Census (No. 3)-Population by Region. 2021. http://www.stats.gov.cn/english/PressRelease/202105/t20210510_1817188.html (accessed August 1 2023).

12. StataCorp. Stata Statistical Software: Release 17. College Station, TX: StataCorp LLC; 2021.

13. Dyer O. Covid-19: Leaked cremation data hint at true scale of China’s death rate. BMJ 2023; 382: p1760.

14. Glanz J, Hvistendahl M, Chang A. How Deadly Was China’s Covid Wave? The New York Times. February 15, 2023.

15. Xiao H, Wang Z, Liu F, Unger JM. Excess All-Cause Mortality in China After Ending the Zero COVID Policy. JAMA Netw Open 2023; 6(8): e2330877.

16. Davidson H. Vaccines are key to China’s zero-Covid exit but scepticism poses challenge. The Guardian. December 2, 2022.

17. Keyes KM, Rutherford C, Popham F, Martins SS, Gray L. How Healthy Are Survey Respondents Compared with the General Population?: Using Survey-linked Death Records to Compare Mortality Outcomes. Epidemiology 2018; 29(2): 299–307.

## Appendix 2 References

1. Main Data of the Seventh National Population Census. 2021 http://www.stats.gov.cn/english/PressRelease/202105/t20210510_1817185.html (accessed August 1 2023).

2. Akimov A, Gemueva K, Semenova N., The Seventh Population Census in the PRC: Results and Prospects of the Country’s Demographic Development. Her Russ Acad Sci 2022; 91: 724-35.

